# Expanding optimization ensemble model methods for forecasting seasonal influenza in the U.S.

**DOI:** 10.1101/2025.11.24.25340597

**Authors:** Benjamin Benteke Longaou, Rhiannon Löster, Pengfei Yue, David Lyver, Christopher M. van Bommel, Edward W. Thommes, Monica-Gabriela Cojocaru

**Affiliations:** University of Guelph, Stone Rd E 50, Guelph, Ontario, Canada; Sanofi, Steeles Ave 1755, North York, Ontario, Canada

**Keywords:** Infectious disease, Modeling, Forecast evaluation, Forecasting

## Abstract

Each year, the seasonal influenza epidemic sees significant variability in its evolution. Accurate forecasts of future influenza cases are important for planning public health responses. The United States Centers for Disease Control and Prevention (CDC) has annually organized the FluSight competition (https://github.com/cdcepi/FluSight-forecast-hub) to solicit forecasts from participating teams over horizons of 1, 2, and 3 weeks ahead. Using these data, the CDC produces an ensemble forecast of all submitted forecasts. In this paper, we introduce a new weight-based ensemble forecasting method to consider predicting laboratory-confirmed influenza hospital admissions for the 2024-2025 season. The method consists of determining optimal weights that are updated week-by-week throughout the FluSight competition to minimize the mean squared error (MSE) of a blend of teams’ previous forecasts compared to the truth data. Using these weights over an expanding time window starting at the beginning of the season (late Fall), we produce our own future forecasts; we call our method the expanding window optimization ensemble (EWO). To improve our method’s performance vis-a-vis the CDC ensemble model, we further introduce the Adjusted-Weights expanding window optimization ensemble (Adw-EWO) method. This new forecast is obtained by adding a correction term to the original EWO forecast, controlled by a parameter *π* ∈ (0, 1). We adaptively tune *π* to enhance forecasting performance of Adw-EWO vis-a-vis the CDC ensemble model. The correction term is computed using only the forecast errors at horizon 0 and is then applied uniformly across all forecast horizons. Our results show that the Adw-EWO method consistently outperforms the EWO across all horizons. Moreover, the Adw-EWO outperforms the CDC ensemble model at horizons 0, 1, and 2, while at horizon 3, the performance of Adw-EWO and the CDC ensemble was roughly comparable.

## 1. Introduction

Forecasting for disease prevalence involves employing predictive models to estimate the future incidence and spread of diseases, which is a critical tool for public health planning and response. Real-time epidemic forecasting can predict geographic disease spread and case counts, aiding public health interventions during outbreaks. However, challenges such as data requirements in epidemic surveillance may arise Desai, Kraemer, Bhatia, Cori, Nouvellet, Herringer, Cohn, Carrion, Brownstein, Madoff and Lassmann (2019).

One disease that faces challenges in forecasting is seasonal influenza. Every year, the seasonal influenza epidemic occurs with large variability in the start of the season and the number of cases throughout the season, making it difficult to anticipate the impact of a season at its beginning. Improving flu forecasting aids in reducing the uncertainty in this variability by providing more accurate predictions of case counts by week and region, which allows for advanced planning in response to predicted changes, such as increased hospitalizations. The Centers for Disease Control and Prevention (CDC) considers several metrics to monitor the current season, including virologic surveillance, outpatient illness surveillance, hospitalization surveillance, and mortality surveillance Centers for Disease Control and Prevention (2024b), but on their own, these metrics do not provide adequate insight into how the flu season will evolve.

To supplement its surveillance data, the CDC launched the “Predict the Influenza Season Challenge” in 2013 Biggerstaff, Alper, Dredze, Fox, Fung, Hickmann, Lewis, Rosenfeld, Shaman, Tsou et al. (2016). This competition was designed to encourage external researchers to forecast the 2013-2014 flu season. When the competition was launched, it was noted that “Mathematical and statistical models can be useful in predicting the timing and impact of the influenza season, but no models published to date have successfully predicted key influenza season milestones with sufficient accuracy.” Centers for Disease Control and Prevention et al. (2013) The goal of hosting the competition was to promote the innovation required to improve the accuracy of influenza forecasts and allow the consideration of multiple techniques from multiple researchers. In subsequent years, the CDC has organized the FluSight competition, and provided data, targets to predict, and evaluation of those predictions, while research teams submit forecasts each week Centers for Disease Control and Prevention (2024a).

Having early insights into the expected beginning, peak, and intensity of the influenza season, rather than observing these aspects as they occur, allows for preparations to be made in anticipation of changes in flu activity. At an individual or community level, knowing anticipated periods of increased flu cases allows for taking measures to reduce contact with others and can be used to encourage vaccination. In terms of the public health response, forecasts can be used to help plan the distribution of antiviral treatments, anticipate sudden influxes in hospitalizations, and therefore the need for health care workers, hospital beds, and treatments. Accurate forecasts help to reduce the costs associated with the disease by more effectively using resources where they are most needed.

Ensemble forecasts, which base their predictions on multiple component models, provide advantages over a single model Bates and Granger (1969); Wolpert (1992); Polikar (2006); Hastie, Tibshirani and Friedman (2009). The idea behind an ensemble forecast is to take a collection of independently determined forecasts and combine the information to form an overarching forecast that reflects the information determined by the component models. The goal is to reduce bias that may be present in a component model due to the particular assumptions that went into its development by balancing them out against other models making different assumptions. Ensemble methods have been successfully utilized in weather forecasting Gneiting and Raftery (2005); Maqsood, Khan and Abraham (2004), and their efficacy has been demonstrated in the prediction of influenza-like illness (ILI) in recent studies De Amorim, Deardon and Saini (2021); Reich, McGowan, Yamana, Tushar, Ray, Osthus, Kandula, Brooks, Crawford-Crudell, Gibson, Moore, Silva, Biggerstaff, Johansson, Rosenfeld and Shaman (2019a); McAndrew and Reich (2021); Ray and Reich (2017). Moreover, McAndrew and Reich McAndrew and Reich (2021) showed that adaptive ensemble models that update weights weekly based on new data can outperform static ensembles that use fixed weights throughout the influenza season.

In this paper, we introduce two novel adaptive ensemble approaches that update using published data from the CDC FluSight competition. Subsequently, the performance of these approaches is compared with existing FluSight ensemble methods.

## 2. Materials and methods

This section describes the FluSight competition, the construction of a general ensemble model, the Expanding Window Optimization (EWO) method, the Adjusted Weighted EWO (Adw-EWO), and the evaluation procedures. We begin by introducing the FluSight competition and its related concepts, followed by a description of the ensemble model, which serves as the foundation of this study. Next, we present our proposed forecasting methods, and finally, we detail the evaluation metrics used to assess model performance.

### 2.1. FluSight competition

As of the 2021-2022 U. S. influenza season, laboratory-confirmed influenza hospital admissions are used as the standard target data for forecasts. The data for 2021-2022, 2022-2023, and 2023-2024 are used as the truth data for the current 2024-2025 influenza season and are available through the FluSight GitHub repository target data CDCEpi (2024). This data is available at the state and national levels, and is also available per facility in which these cases are confirmed. Data for each team’s forecast submissions, including the CDC ensemble forecast, was also made available through the FluSight GitHub repository CDCEpi (2024).

For the 2024-2025 influenza season, the CDC FluSight competition requested participating teams to submit weekly forecasts from November 20, 2024 to May 31, 2025. Weeks are defined in terms of epidemiological weeks (EW or Epi Week), and are counted for each calendar year with the week, Sunday to Saturday, containing January 1^st^ labeled EW 1 Centers for Disease Control and Prevention (2023). Participating teams were invited to submit their forecasts to FluSight by 11:00 PM Eastern Time each Wednesday. Each submission is associated with the epidemiological week (EW) that ends on Saturday following the submission deadline, which serves as the reference date for the prediction. The forecasts requested are quantile predictions for laboratory confirmed influenza hospital admissions at the state level and the national level of the United States during subsequent weeks and probability predictions for the direction (“large increase”, “increase”, “stable”, “decrease”, “large decrease”) of the trajectory of hospitalization rates. The requested quantiles are: {0.01, 0.025: 0.05: 0.975^1^, 0.99.} The subsequent weekly targets are referred to as “horizons”, with values −1, 0, 1, 2, and 3, corresponding to the week before −1), the current week (0), the next week (1), two weeks ahead (2), and three weeks ahead (3), respectively. Note that, in a typical competition year, the forecasts are produced and evaluated (for performance) on horizons 0–3 only, as observed true hospitalization values are available for horizon −1 before the teams’ weekly submission deadline (approximately noon on Wednesdays). The Target End Date for each forecast is defined as the reference date plus horizon × 7 days. Further details on the 2023-2024 and 2024-2025 FluSight competitions can be found in the FluSight GitHub repository CDCEpi (2024).

The FluSight Ensemble model is produced by taking the median value corresponding to each quantile among all eligible forecasts, as described in Mathis, Webber, León, Murray, Sun, White, Brooks, Green, Hu, Rosenfeld et al. (2024). The FluSight Baseline forecast is computed by taking the previous week’s value as the median of the prediction interval, with the variability of the distribution for each jurisdiction generated based on the positive and negative differences between consecutive weeks of all prior observations, truncated to prevent negative values. An overview of the Baseline method is provided in Mathis et al. (2024), and further details about its implementation can be found in Cramer, Ray, Lopez, Bracher, Brennen, Castro Rivadeneira, Gerding, Gneiting, House, Huang et al. (2022b); Cramer, Huang, Wang, Ray, Cornell, Bracher, Brennen, Rivadeneira, Gerding, House et al. (2022a).

### 2.2. General Ensemble construction method

Let us denote by *T* is the total number of weeks within a typical season for forecasting hospitalizations due to influenza. For instance, *T* is 23, 32, 30, and 27 for 2021-2022, 2022-2023, 2023-2024, and 2024-2025, respectively. Let us further denote by *w* a typical week in a given season. Further, let *M* refer to the total number of consistent^2^ teams in a season, while *h* will denote the index of forecast horizons (see Table 1 below):

**Table 1.**
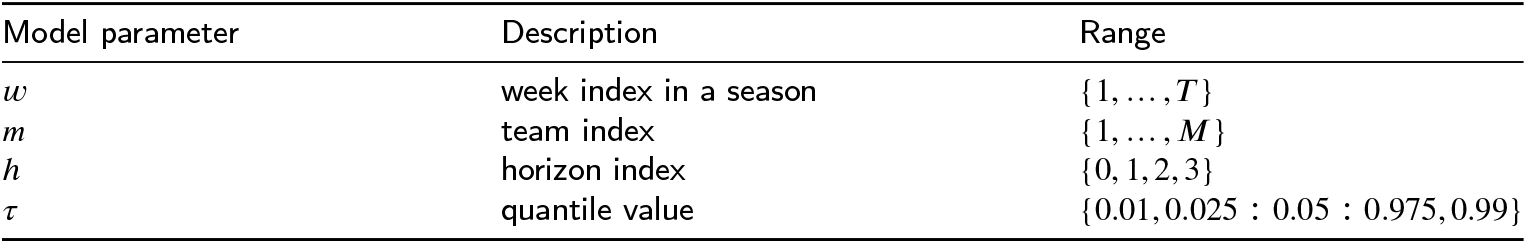
Model parameters, descriptions, and ranges.

| Model parameter | Description | Range |
| --- | --- | --- |
| $w$ | week index in a season | $\{1, \dots, T\}$ |
| $m$ | team index | $\{1, \dots, M\}$ |
| $h$ | horizon index | $\{0, 1, 2, 3\}$ |
| $\tau$ | quantile value | $\{0.01, 0.025 : 0.05 : 0.975, 0.99\}$ |

Let 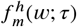 be a quantile forecast submitted in week *w* for participant team *m* for a horizon *h*, and a given specific quantile *τ*. The goal of the ensemble method is to blend teams’ quantile forecasts, such that

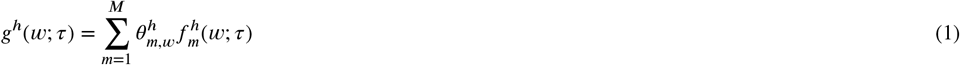

is better than any single 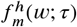 in the ensemble over horizons. The goal of our method is to estimate the weights 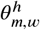 via a constrained optimization problem so that all weights are non-negative and sum up to 1. However, since it is not possible to observe the quantiles and instead only to observe confirmed influenza hospital admissions, we use the point forecast of teams, denoted by

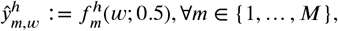

and the truth data for the corresponding week *w*, denoted *y*_*w*_, to perform our optimization. Therefore, for each fixed week *w* and each fixed horizon *h*, our optimization problem becomes:

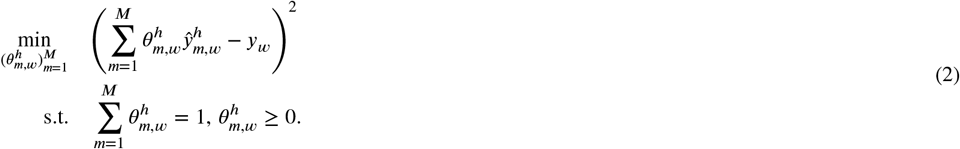

The problem (2) is only for one single week. We further use a supervised machine learning framework that involves collecting forecast data for all teams {1, …, *M*}, from different weeks *w*, as well as the corresponding truth data *y*_*w*_, and formulate a learning problem to estimate new weight values 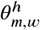 every subsequent week ofteh season. Due to the dependency of data between weeks in a season (as they are time series data) using them without leveraging their structure may lead to non-optimal weights during learning. In this paper, we use an expanding time window to organize our teams’ weekly data before passing it to our learning algorithm. The goal of learning every week is to find optimal weights for each team and use this to blend future quantile predictions of the teams as described in (1); we refer to this method as the expanding window optimization (EWO) method.

### 2.3. Expanding Window Optimization

The expanding window approach enables us to determine the optimal weights for each consistent team by leveraging their historical forecasts from previous seasons and their forecasts from the current season up to the present week. For the 2024–2025 season, we incorporated teams’ historical forecasts from the 2021–2022, 2022–2023, and 2023–2024 seasons, along with their forecasts from the current 2024–2025 season, up to a current week, *w*. A diagram of the basic expanding window technique is shown in Fig. 1, where the left end of the window is fixed to the beginning of the sample, while the right side grows further along the sample size; the horizon refers to the forecasting window. In our case, the expanding window is applied only to the current season and starts from *w* > 4, which helps us to gather observed data for all horizons at teh start of the forecasting season. For example, if *w* = 5, then {*y*_1_, *y*_2_, *y*_3_, *y*_4_}, {*y*_2_, *y*_3_, *y*_4_}, {*y*_3_, *y*_4_}, and {*y*_4_} are the available observed hospitalization data needed to forecast each horizon 0, 1, 2, and 3, respectively.

**Figure 1.**
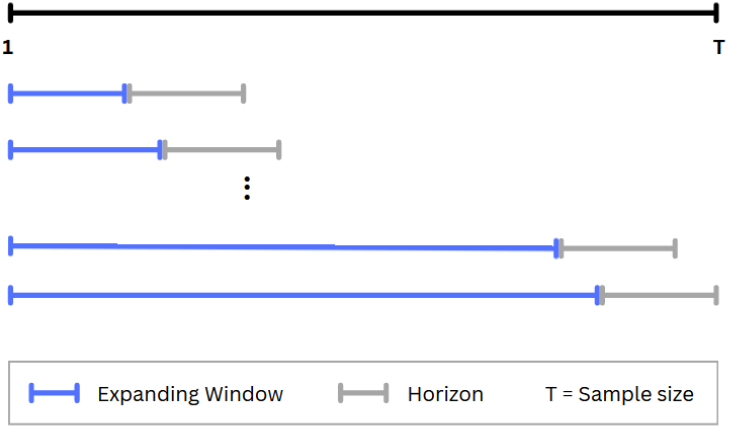
Diagram of expanding window methodology.

Consider now the quantile forecast *f*^*h*^(*w* + *h*; *τ*) submitted in week *w* at horizon *h*, to predict week *w* + *h* (with *τ* denoting the different quantile levels, as before). We determine the weights, denoted 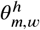, for a given horizon and target date assigned to each team in our ensemble by minimizing the mean square error of the ensemble point (median) forecast with the observed data. This minimization is done on the expanding window [*h* + 1, *w*− 1] alongside the data from the previous seasons (forecasts and observed data). For example, if current week is *w* = 10, then data to be used for forecasting in each horizon will be drawn from weeks: {1, …, *w* − 1 = 9} for horizon *h* = 0, {2, …, *w* − 1 = 9} for horizon *h* = 1, {3, …, *w* − 1 = 9} for horizon *h* = 2, and {4, …, *w* − 1 = 9} for horizon 3. Starting with *w* = 5, for each subsequent submission week *w* > 4 in the current season, the set of weeks within from which data is drawn will expand by one additional week on the right. The optimization problem for a forecast of week *w* at horizon *h* is done using the past seasons data and all the data within the expanding window, and is shown in the equation below:

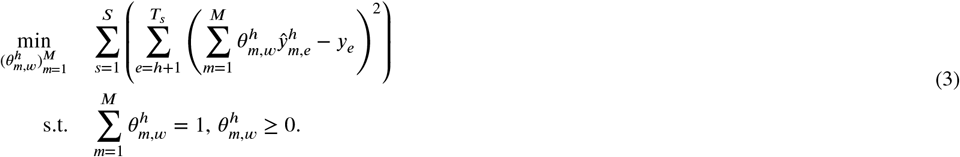

where *S* is the total number of past seasons data, *T*_*s*_ is the total number of weeks in any past seasons: *s* = 1, …, *S* − 1, whereas *T*_*s*_ ≔ *w* is the current week in the current forecasting season.

Let us denote by 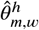 the learned weight for a team *m* and horizon *h* as of week *w*. To learn these teams’ weights, for each horizon *h* and submission week *w*, we solve the optimization problem defined in (3). To this end, we collect the point forecasts 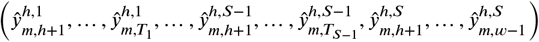 for *m* = 1, …, *M*, along with their corresponding observed data: 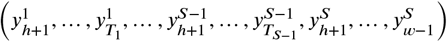. For simplicity, we omit the season index *s* = 1, …, *S* in both the forecasts and observed data in the remainder of the notation. During training, we use forecasts and observed data from week *h* + 1 of the first season up to week *w* − 2 of the current season to learn the weight vector 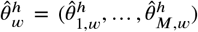. We then validate the learned weights 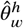 using forecasts and observed values from week *w* − 1, selecting the weight vector that minimizes the validation loss: 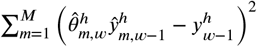, where 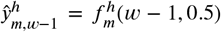. For example, in week 5 of the current season and for horizon *h* = 5, we train using past seasons’ data up to week 4, validate on week 5, and forecast for week 7. Once the optimal weights 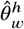 are obtained, we blend the forecasts 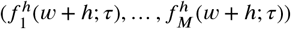 as below: The ensemble model, *g*_*h*_(*w* + *h*; *τ*), is then defined as follows:

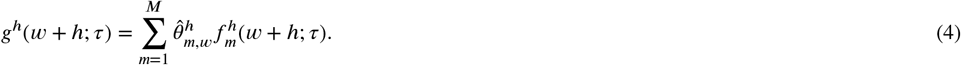

As the learning algorithm, we employ Adam (Adaptive Moment Estimation) Kingma (2014) optimizer with a learning rate of 10^−3^, *β*_1_ = 0.9, and *β*_2_ = 0.999 to solve (3), following the configuration suggested in the original paper. To ensure convergence to an optimal solution 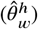, we apply early stopping based on the validation loss—training stops when no further improvement is observed.

### 2.4. Adjusted weighted-EWO

The Adjusted weighted-EWO (Adw-EWO) aims to improve the forecast performance of the EWO ensemble by adding a correction term to the forecast generated by EWO for all horizons. The correction term is computed using only the forecast errors estimated at horizon 0 and then applied uniformly across all forecast horizons. To compute a needed correction we introduce a short term memory term that adjusts the EWO forecasts. We consider three scenarios: one week behind, two weeks behind, and three weeks behind. As before, in the 2024–2025 season forecasting began in week *w* = 5, with adjustments applied from week *w* = 6 onward.

In the one-week memory scenario, assuming a current week *w*^*^, the correction term is computed from the EWO forecast error (w.r.t. true data) of the previous week (*w*^*^ − 1), and scaled by a discount factor *π*, which determines the strength of the correction. The two-week memory scenario incorporates EWO errors (w.r.t true data) from the past two weeks (*w*^*^ − 1, *w*^*^ − 2), and uses a weighted average: the most recent error (from week *w*^*^ − 1) is weighted by *π* and the earlier error (from week *w*^*^ − 2) by *π*^2^, ensuring more emphasis to recent error. Similarly, the three-week memory correction includes errors from the previous three weeks, applying weights of *π, π*^2^, and *π*^3^, respectively. Starting from week 6, each subsequent forecast is corrected by shifting it toward the corresponding correction term. In addition, *π* needs to be tuned effectively (from a uniform distribution of values in 0, 1) - see 6 below). The following expression illustrates the case of computing an optimal discount factor *π* for the one-week memory correction^3^:

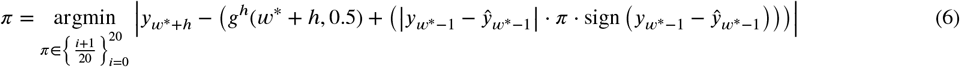

We note that in Equation 6, we use the observed data at *w*^*^ + *h* for *h* = 0 and *τ* = 0.5. We then compute the correction term and apply it to forecasts at horizons *h* = 0, 1, 2, 3 for all *τ*. Therefore, it is reasonable to also include comparisons of our proposed methods with the CDC ensemble at horizons 1, 2, and 3. Note that once the forecast for week *w*^*^ is corrected, the corrected value serves as the basis for correcting future weeks. The pseudocode detailing our proposed methods is presented in the Appendix.

### 2.5. Evaluation methods

For the current 2023-2024 influenza season, the scoring method proposed by the CDC is weighted interval score (WIS). The WIS generates interval scores for probabilistic forecasts expressed in terms of quantiles, and therefore, accounts for dispersion, under and over prediction Mathis et al. (2024). It is composed of a simple absolute difference score for the median quantile (point forecast estimate), score values for the width of each interval, and penalties for intervals that do not contain the true value. A lower absolute WIS value is attributed to more accurate forecasts compared to a higher absolute WIS value. Additionally, the mean absolute error (MAE) value was also reported.

#### 2.5.1. Weighted Interval Score (WIS)

For a given prediction interval (1 − *τ*) × 100% of a model’s forecast *F*, the interval score is defined as

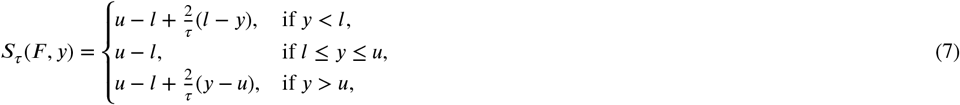

where *u* (*l*) represents the upper (lower) limit of the prediction interval and *y* is the actual true value. The WIS generalizes the interval score to multiple prediction intervals and is defined as:

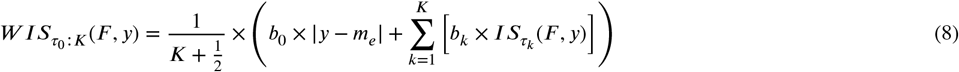

Here, *K* denotes the total number of intervals considered, *m*_*e*_ is the forecast median, and *w*_*k*_ are the non-negative weights assigned to the different intervals. Following a standard approach, we set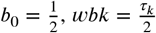, and consider 23 prediction intervals (*τ*_*k*_ = 0.01, 0.025, 0.05, 0.1, 0.15, 0.2, 0.25, 0.3, 0.35, 0.4, 0.45, 0.5, 0.55, 0.6, 0.65, 0.7, 0.75, 0.8, 0.85, 0.9, 0.95, 0.975, 0.99).

For each horizon *h*, the **Mean Weighted Interval Score (MWIS)** is the average of the WIS across weeks, and the smaller values indicate better forecast accuracy.

#### 2.5.2. Mean absolute error (MAE)

Given a median (point) forecast *ŷ* and a corresponding actual value *y*, the mean absolute error is simply calculated as: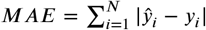. In our evaluation, for each horizon *h*, the MAE is the average of the absolute error over weeks, and smaller values indicate better forecast accuracy.

## 3. Results and discussions

In this section, we run EWO in a real-time setting for each week *w* > 4 of the 2024–2025 season, updating forecasts weekly using historical data of consistent teams from the 2021–2022, 2022–2023, and 2023–2024 seasons, as well as all available weeks from the 2024–2025 season up to week *w* − 1 (depending on the forecast horizon *h*). Next, we apply Adw-EWO to correct the forecasts generated by EWO.

Our analysis includes predictions at all horizons (0, 1, 2, and 3). Then, for each horizon, we compute the Mean Absolute Error (MAE) and the Mean Weighted Interval Score (MWIS) for EWO, Adw-EWO (one-week, two-week, and three-week correction scenarios), and the CDC Ensemble during the 2024–2025 season. Specifically, we compare models in three ways: per horizon, by aggregating MAE and WIS across all horizons, and by aggregating MAE and WIS across horizons 1, 2, and 3. We also discuss the corrections, especially the number of past weeks to keep in memory to compute the correction term. Finally, we analyze the weekly WIS values of our proposed method (EWO) and examine the team weights produced by EWO throughout the 2024–2025 season.

Since no forecasts were submitted on 2025-01-25, we missed the forecasts corresponding to the target dates or epidemiological week (EW) 2025-01-25, 2025-02-01, 2025-02-08, and 2025-02-15, which align with horizons 0, 1, 2, and 3, respectively. Forecasting with EWO begins at week index 5. Corrections start at week indices 6, 7, and 8 for the one-week-behind, two-week-behind, and three-week-behind scenarios, respectively. Let us name forecasting weeks: *w*_1_= 2024-11-23, *w*_2_= 2024-11-30, *w*_3_= 2024-12-07, *w*_4_= 2024-12-14, *w*_5_= 2024-12-21, *w*_6_= 2024-12-28, *w*_7_= 2025-01-04, *w*_8_= 2025-01-11, *w*_9_= 2025-01-18, *w*_10_= 2025-01-25, *w*_11_= 2025-02-01, *w*_12_= 2025-02-08, and so on.

First, Tables 2, 3, 4, and 5 show that the Adw-EWO, across all scenarios and horizons, outperformed the EWO in both MAE and MWIS, which aligns with our expectations. Furthermore, for horizons 0, 1, and 2, the Adw-EWO (one-week behind) significantly outperformed the CDC Ensemble in terms of MAE and MWIS, with only a small performance gap in MWIS at horizon 2. However, at horizon 3, the CDC Ensemble demonstrated a lower MAE compared to the Adw-EWO, while the Adw-EWO (two-week behind) achieved the lowest MWIS among the models, outperforming the CDC Ensemble in that metric.

**Table 2.** Performance Metrics for Different Models (Horizon = 0)

| Model | MAE | Mean WIS |
| --- | --- | --- |
| CDC Ensemble | 3489.31 | 2630.17 |
| Uncorrected EWO | 3387.01 | 2440.30 |
| Adw-EWO (One-week behind with various $\pi$) | 2113.71 | 1660.32 |
| Adw-EWO (Two-week behind with various $\pi$) | 2401.81 | 1874.30 |
| Adw-EWO (Three-week behind with various $\pi$) | 2379.60 | 1868.95 |

**Table 3.** Performance Metrics for Different Models (Horizon = 1)

| Model | MAE | Mean WIS |
| --- | --- | --- |
| CDC Ensemble | 4919.14 | 3693.90 |
| Uncorrected EWO | 5484.60 | 3833.74 |
| Adw-EWO (One-week behind with various $\pi$) | 4671.57 | 3368.01 |
| Adw-EWO (Two-week behind with various $\pi$) | 5008.51 | 3582.70 |
| Adw-EWO (Three-week behind with various $\pi$) | 4983.66 | 3542.84 |

**Table 4.**
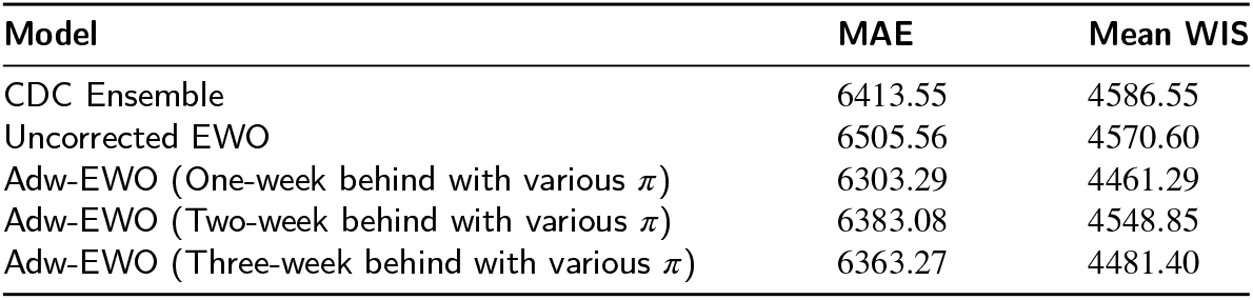
Performance Metrics for Different Models (Horizon = 2)

**Table 5.** Performance Metrics for Different Models (Horizon = 3)

| Model | MAE | Mean WIS |
| --- | --- | --- |
| CDC Ensemble | 8259.57 | 5648.80 |
| Uncorrected EWO | 8999.51 | 6051.87 |
| Adw-EWO (One-week behind with various $\pi$) | 8546.95 | 5858.87 |
| Adw-EWO (Two-week behind with various $\pi$) | 8420.92 | 5129.60 |
| Adw-EWO (Three-week behind with various $\pi$) | 8394.70 | 5640.91 |

Table 5 shows that the one-week-behind version of Adw-EWO did not perform well at horizon 3, whereas the two-week and three-week variants achieved lower MWIS values in comparison. A similar trend can be observed at horizon 2, where the different versions of Adw-EWO produced comparable MAE and MWIS values. These results suggest that incorporating more past weeks into memory becomes increasingly beneficial as the forecasting horizon extends.

Overall, Table 6 shows that all versions of our Adw-EWO model (one-week, two-week, and three-week) outperformed the CDC Ensemble.

**Table 6.**
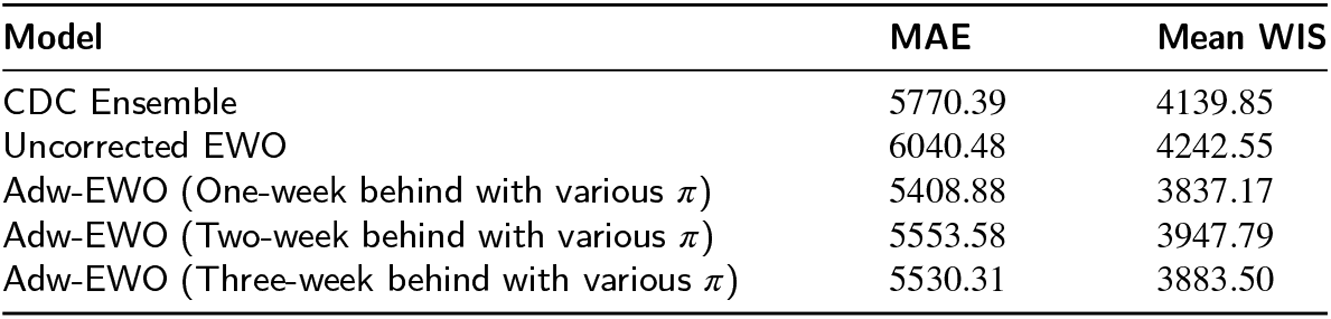
Mean of Performance Metrics for Different Models across horizons.

| Model | MAE | Mean WIS |
| --- | --- | --- |
| CDC Ensemble | 5770.39 | 4139.85 |
| Uncorrected EWO | 6040.48 | 4242.55 |
| Adw-EWO (One-week behind with various $\pi$) | 5408.88 | 3837.17 |
| Adw-EWO (Two-week behind with various $\pi$) | 5553.58 | 3947.79 |
| Adw-EWO (Three-week behind with various $\pi$) | 5530.31 | 3883.50 |

Table 7 shows that all versions of our Adw-EWO model (one-week and three-week) outperformed the CDC Ensemble regarding WIS, when excluding horizon 0.

**Table 7.** Mean of Performance Metrics for Different Models across horizons (h= 0 excluded)

| Model | MAE | Mean WIS |
| --- | --- | --- |
| CDC Ensemble | 6530.75 | 4643.08 |
| Uncorrected EWO | 6898.37 | 4818.74 |
| Adw-EWO (One-week behind with various $\pi$) | 6507.27 | 4562.79 |
| Adw-EWO (Two-week behind with various $\pi$) | 6604.17 | 4638.96 |
| Adw-EWO (Three-week behind with various $\pi$) | 6580.54 | 4555.05 |

The tables above provide the aggregated mean absolute error (MAE) and weighted interval score (WIS) values, and give an overall performance of the models. Let us now discuss the weekly WIS values individually. The Fig 2 shows that the peak week is 2025-02-08. To discuss the weekly WIS values, we divide the epidemic curve into three parts: the growth phase, around the peak, and the decay phase. The growth phase (from 2024-12-21 to 2025-01-11) is around the beginning to three weeks before the peak, around the peak (from 2025-01-18 to 2025-02-22) is from two weeks before the peak and two weeks after the peak, and the decay phase (from 2025-03-01 to the end of the season) is three weeks after the peak to the end of the season.

**Figure 2.**
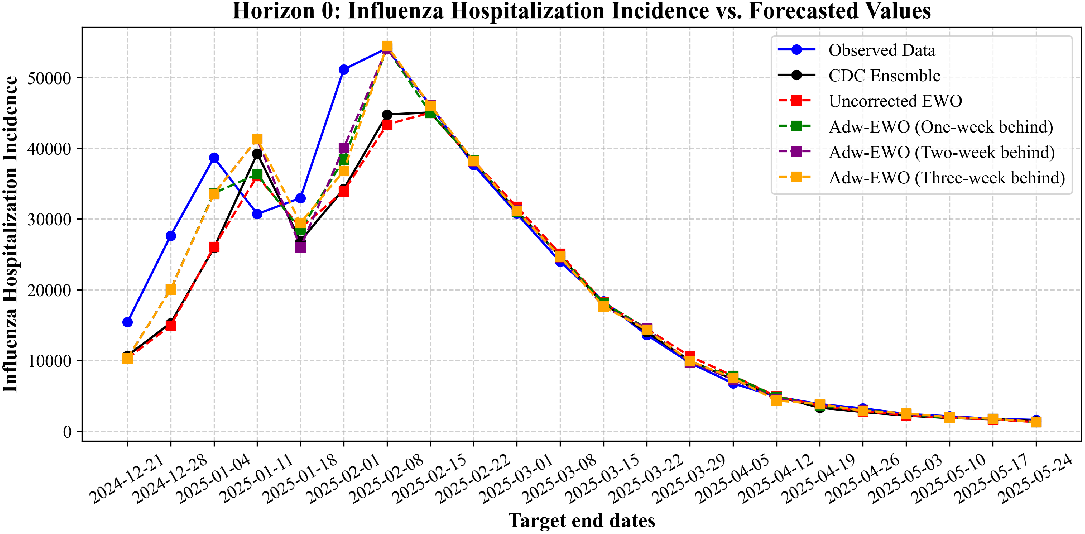
Horizon 0: FluSight competition 2024-2025.

Fig 3 shows that at horizon 0, the Adw-EWO models (one-week, two-week, and three-week) performed better than the CDC Ensemble during the growth phase and around the peak. However, both Adw-EWO and the CDC Ensemble performed similarly well in the decay phase.

**Figure 3.**
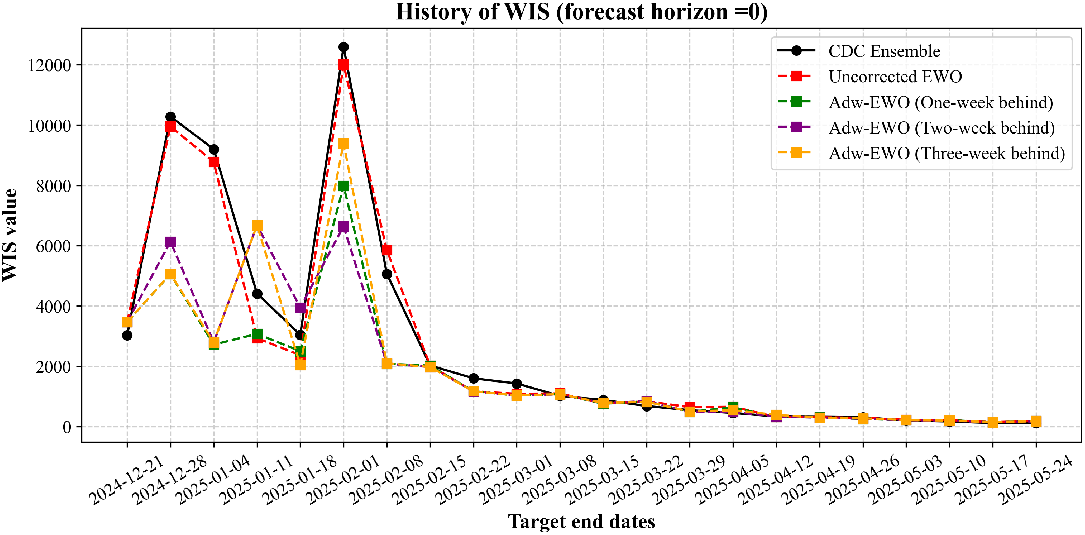
WIS: History for each team (forecast horizon = 0).

Fig 4 shows that at horizon 1, the Adw-EWO models and the CDC Ensemble performed almost equally during the growth phase, while Adw-EWO outperformed the CDC Ensemble around the peak. In the decay phase, however, the CDC Ensemble outperformed Adw-EWO.

**Figure 4.**
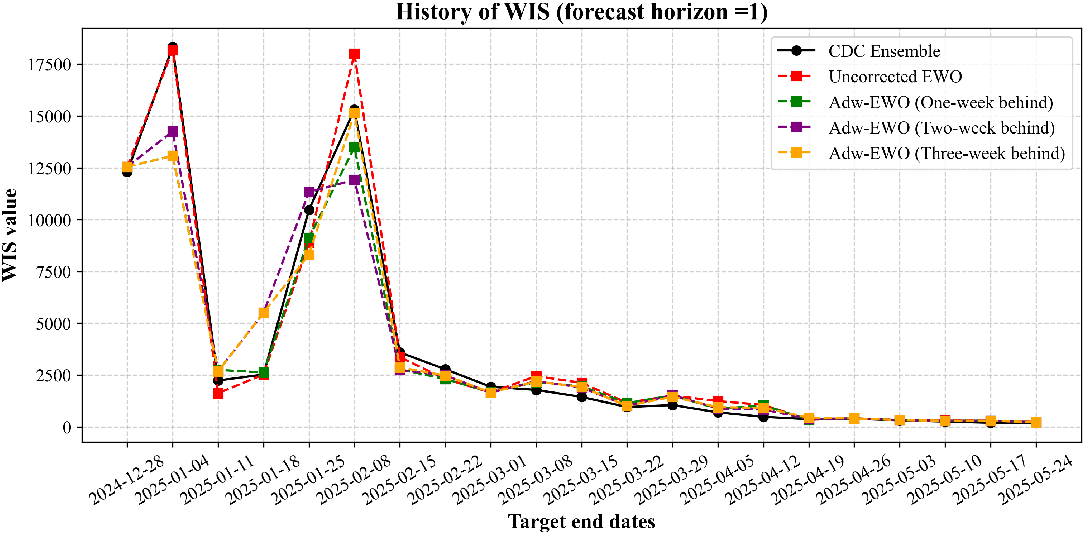
WIS: History for each team (forecast horizon = 1).

Fig 5 shows that at horizon 2, the Adw-EWO models outperformed the CDC Ensemble during the growth phase and around the peak. Similar to horizon 1, the CDC Ensemble showed better performance than Adw-EWO in the decay phase.

**Figure 5.**
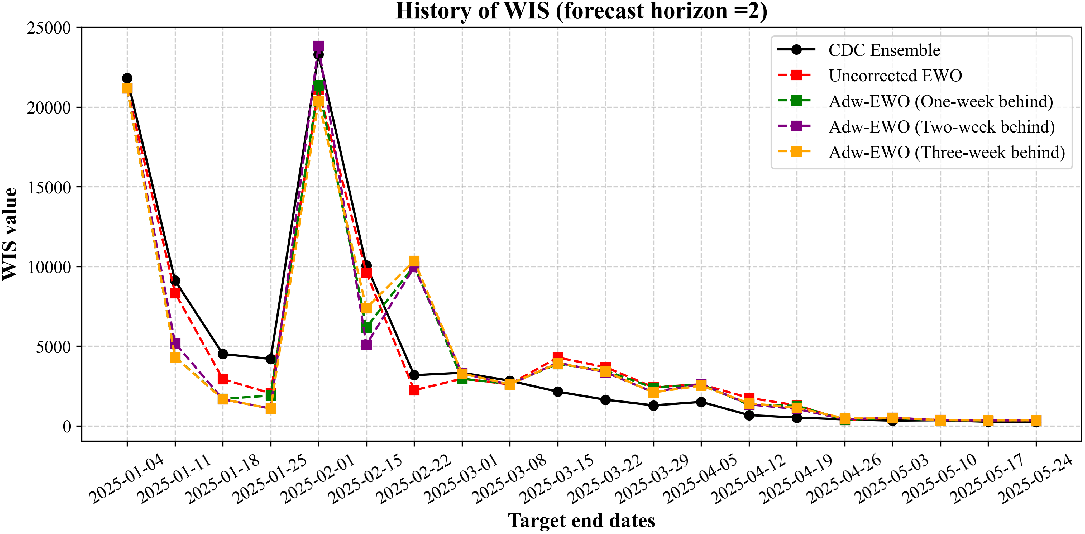
WIS: History for each team (forecast horizon = 2).

Fig 6 shows that at horizon 3, the Adw-EWO models again outperformed the CDC Ensemble during the growth phase and around the peak. Finally, the CDC Ensemble outperformed Adw-EWO in the decay phase.

**Figure 6.**
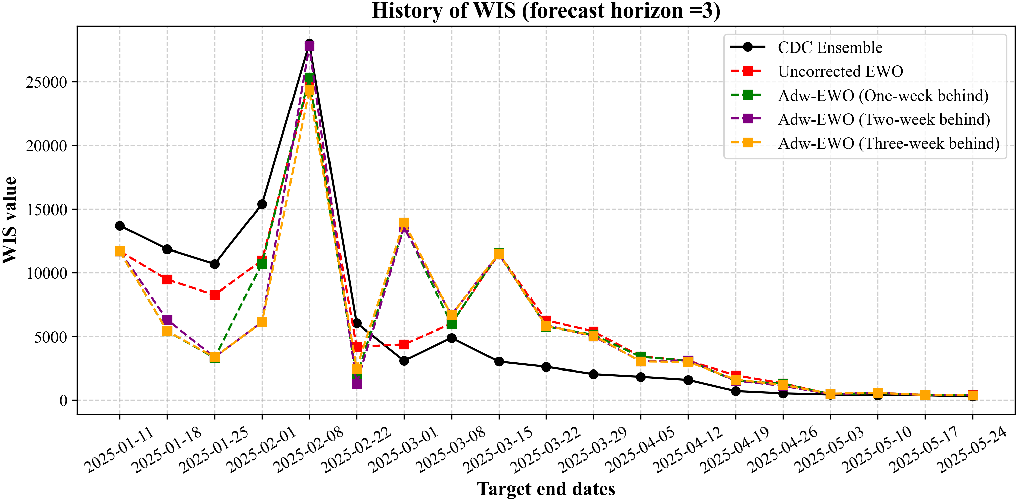
WIS: History for each team (forecast horizon = 3).

Let us now discuss the weights of the teams that contributed to the EWO results. Fig 7 shows that at horizon 0, in the growth phase, teams were assigned almost equal weight. On 2024-12-28, only one team had a high weight, while the others were assigned almost equal weight. On 2024-01-04, three teams had similarly high weights, while the others maintained relatively equal but smaller weights. In the following weeks, the teams were assigned nearly equal weight overall.

**Figure 7.**
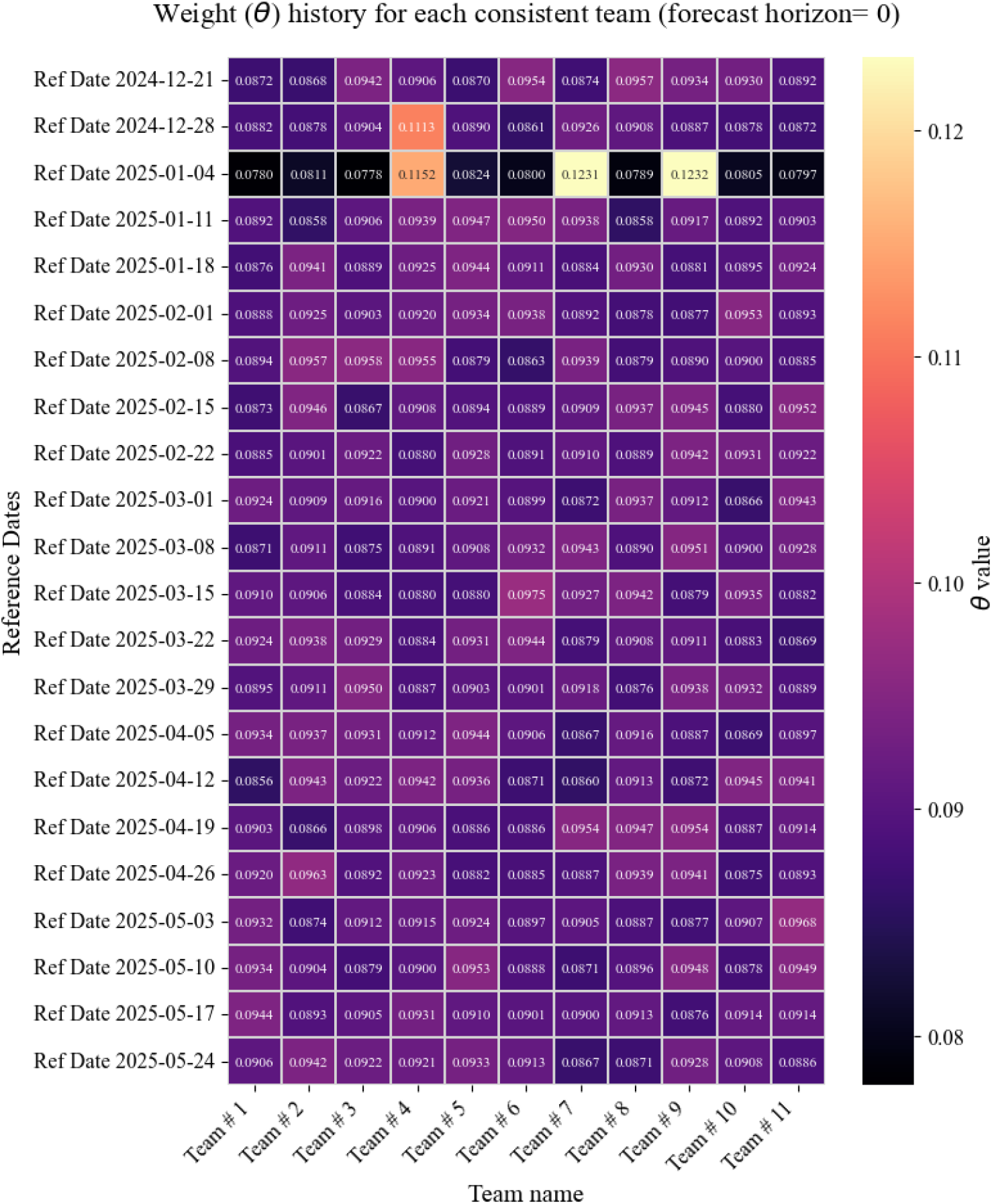
Horizon 0: Weights history for consistent teams in EWO.

Fig 8 shows that at horizon 1, the team weights were almost equal. However, as the peak week approached, the weights began to shift, with some teams receiving higher weights and others lower. In the following weeks (after the peak week), the teams were assigned nearly equal weight overall. This indicates that forecasting the peak in the previous week (horizon 1) was not a particularly difficult task for the teams.

**Figure 8.**
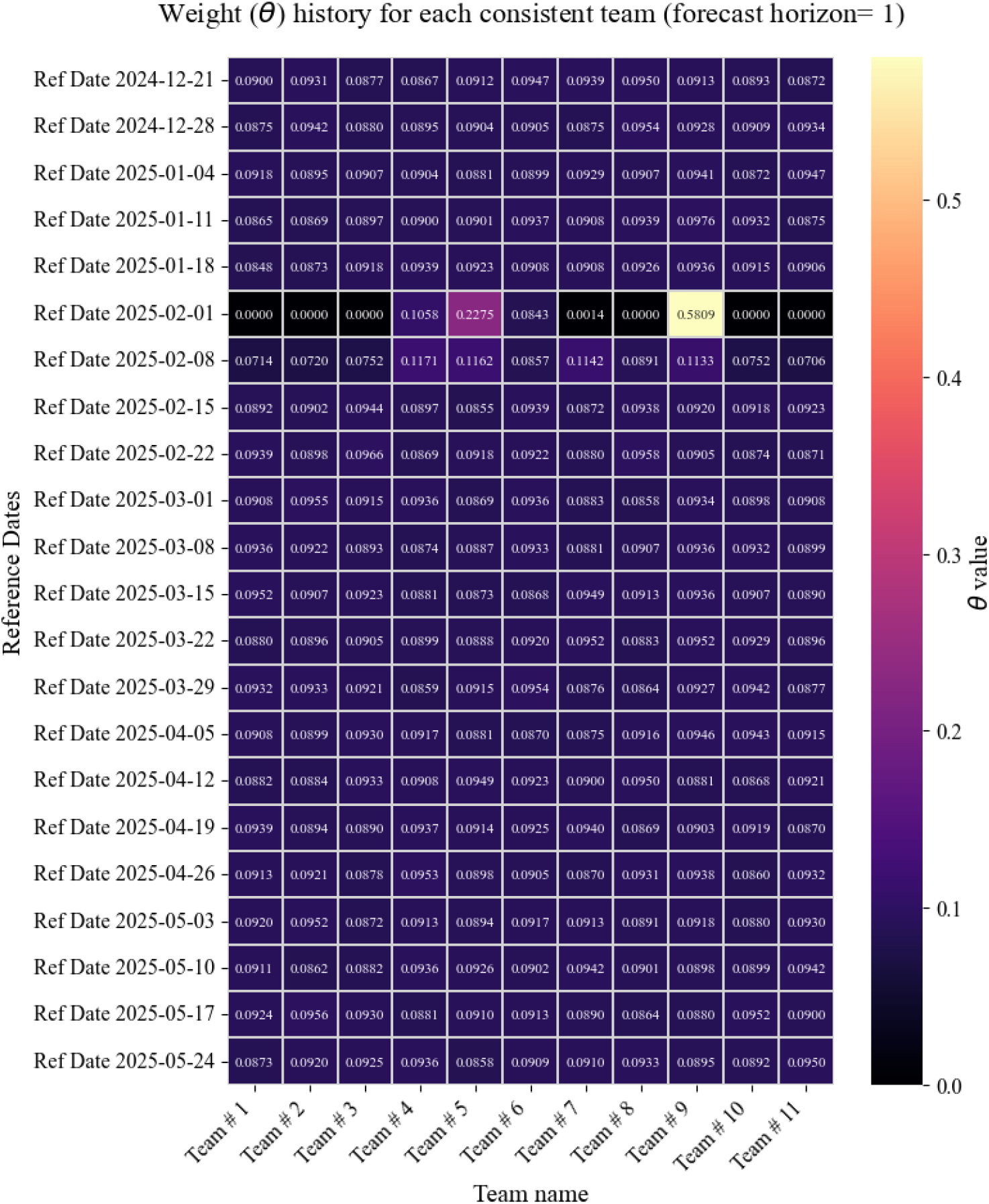
Horizon 1: Weights history for consistent teams in EWO.

Figure 9 shows that at horizon 2, all teams were assigned almost equal weight except on 2025-01-11 and 2025-02-08. On 2025-02-08, we observed that one team was assigned a significantly higher weight (0.6114), demonstrating how the EWO became selective around the peak.

**Figure 9.**
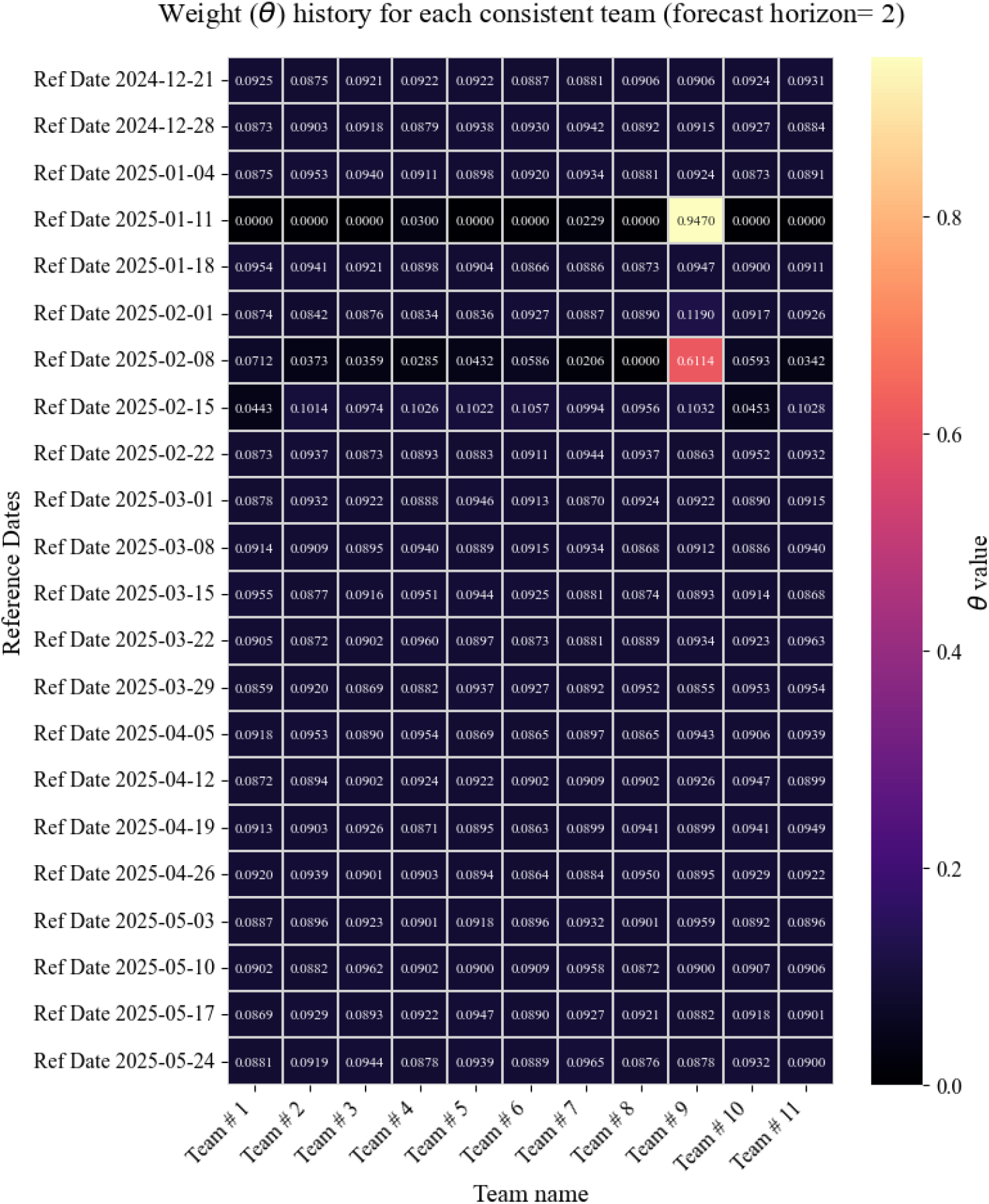
Horizon 2: Weights history for consistent teams in EWO.

Fig 10 shows that at horizon 3, team weights were mostly equal, except around the peak, where the EWO selectively assigned higher weights to certain teams. This highlights the selective behavior of the EWO during around peak phase. The behavior of the team weights suggests that, most of the time, the EWO assigned nearly equal weights to all teams, except during specific periods, such as around the peak week. This limited selectivity may explain why it was unable to consistently outperform the CDC Ensemble.

**Figure 10.**
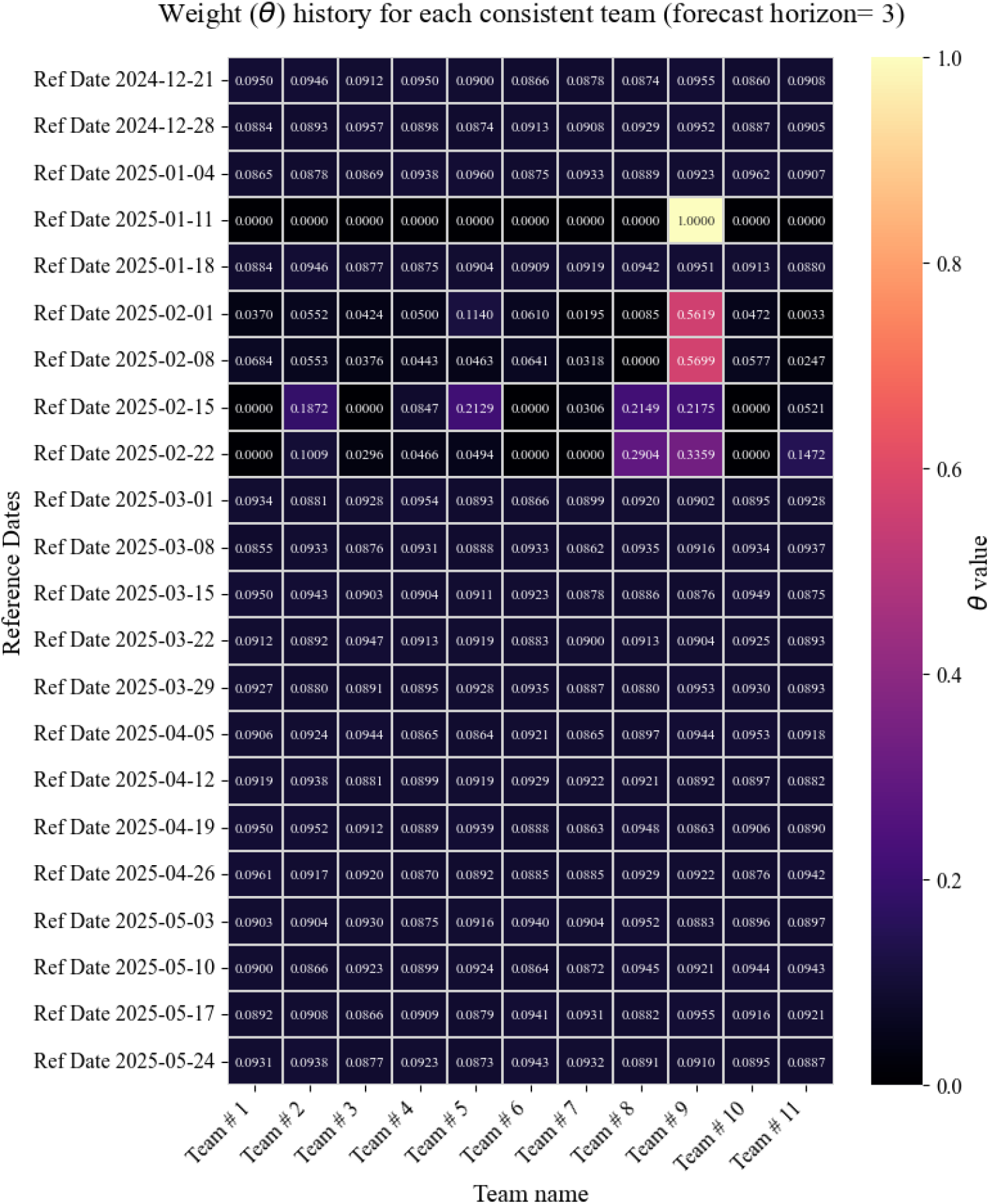
Horizon 3: Weights history for consistent teams in EWO.

## 4. Conclusion

In this paper, we introduced two novel adaptive ensemble approaches to produce forecasts for the CDC FluSight competition. The first, Expanding Window Optimization (EWO), uses the previous forecasts of participating teams to determine optimal weights with which to take a weighted average of team forecasts to produce an ensemble forecast. The second, adjusted-weighted EWO (adw-EWO), applies an additive correction based on the previous performance of the ensemble forecast, with a varying discount factor applied to the correction. On its own, the EWO method is not sufficient to improve on the FluSight Ensemble forecasts, but the adw-EWO method offers an improvement for horizons 0, 1, and 2 at particular values of *π*, and with additional insight into how the optimal value of *π* changes throughout the season, has the potential to be an improvement on the FluSight Ensemble model. Notble are the better performance scores obtained over all horizons by adw-EWO in the exponential growth regimes and around the peak regimes of the season. As with ensemble models, however, identifying time frames in advance where this shift in behaviour occurs is challenging Mathis et al. (2024); McGowan, Biggerstaff, Johansson, Apfeldorf, Ben-Nun, Brooks, Convertino, Erraguntla, Farrow, Freeze et al. (2019); Reich, Brooks, Fox, Kandula, McGowan, Moore, Osthus, Ray, Tushar, Yamana et al. (2019b). Moving forward, previous seasons can offer insight into the general trends and help inform the choice of parameter *π* in the adw-EWO model. Moreover, an analysis of the trade-off between including more memory for corrections versus performance gains can be undertaken.

## Data Availability

All data produced are available online at https://github.com/cdcepi/FluSight-forecast-hub.

## A. Appendix

### Pseudocodes

We include below the detailed description of our EWO method, from an algorithmic viewpoint. Rewrite it to Adam, and explain this choice.

## CRediT authorship contribution statement

**Benjamin Benteke Longaou:** Methodology, Coding, Data curation, Writing. **Rhiannon Löster:** Data curation, Literature Review, Data Visualization. **Pengfei Yue:** Data curation. **David Lyver:** Data curation, visualization. **Christopher M. van Bommel:** Conceptualization, Data curation, Writing, Editing, Proof-reading Code. **Edward W. Thommes:** Data curation, Writing, editing. **Monica-Gabriela Cojocaru:** Conceptualization, Corrected method, Writing, editing.

## Algorithm 1

Adam with Early stopping to determine weight assigned to each team as of week *w* > 4 for horizon *h*

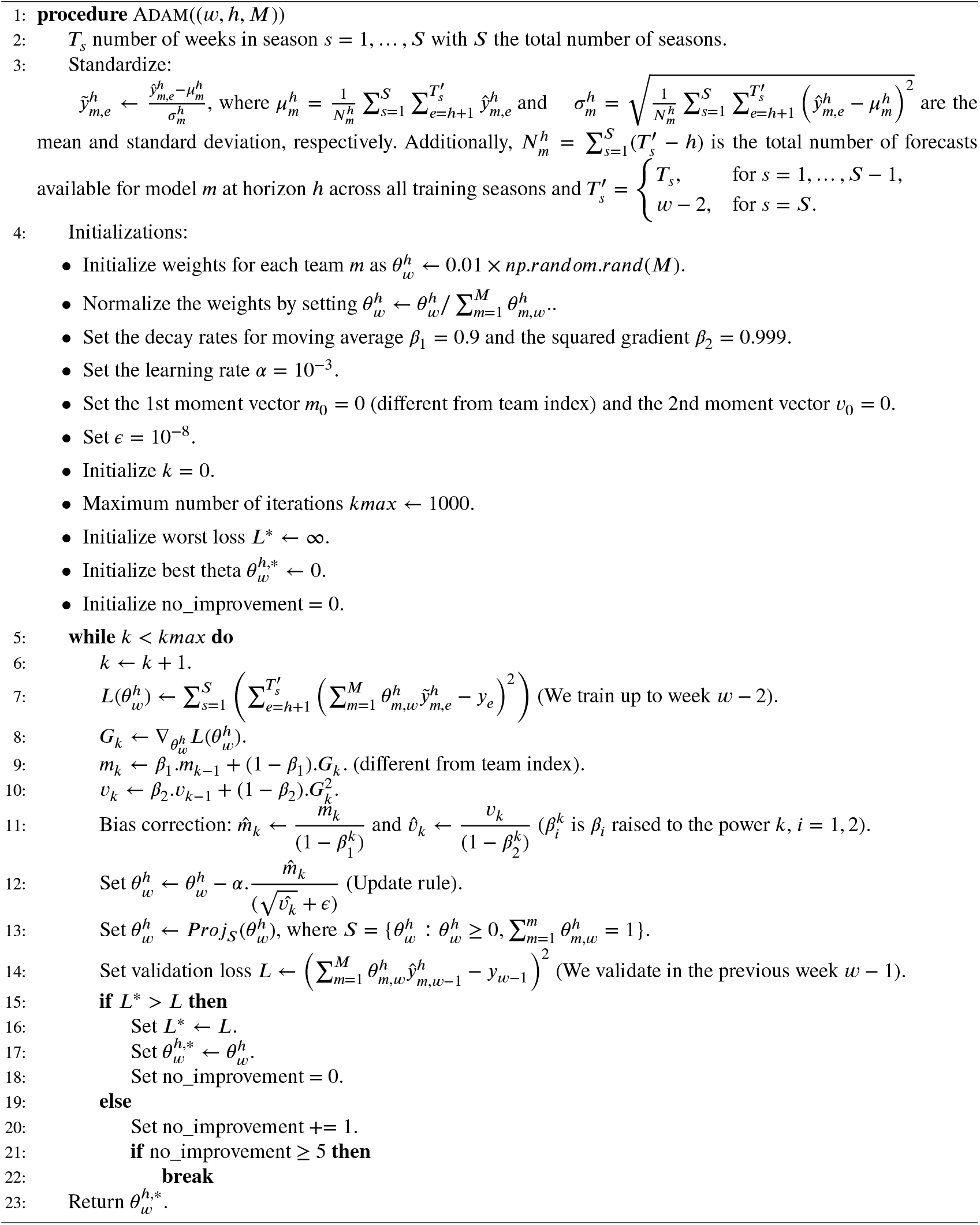

## Algorithm 2

EWO to Produce the (Uncorrected) Forecast Submitted in week *w* at Horizon *h*

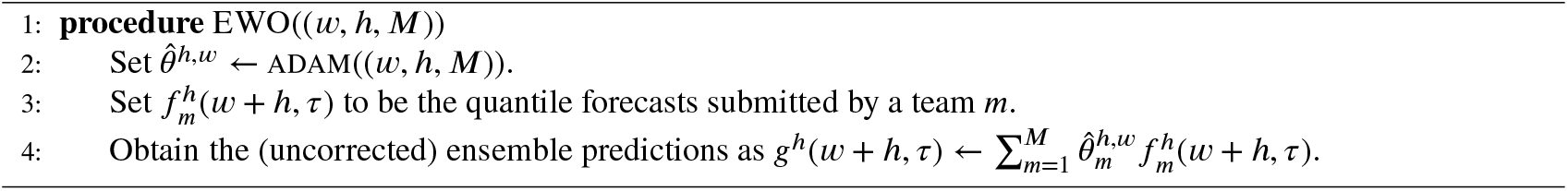

## Algorithm 3

Adjusted-Weighted (Adw) EWO to correct forecast Submitted in week *w*^*^ at horizon *h* with discount factor *π*

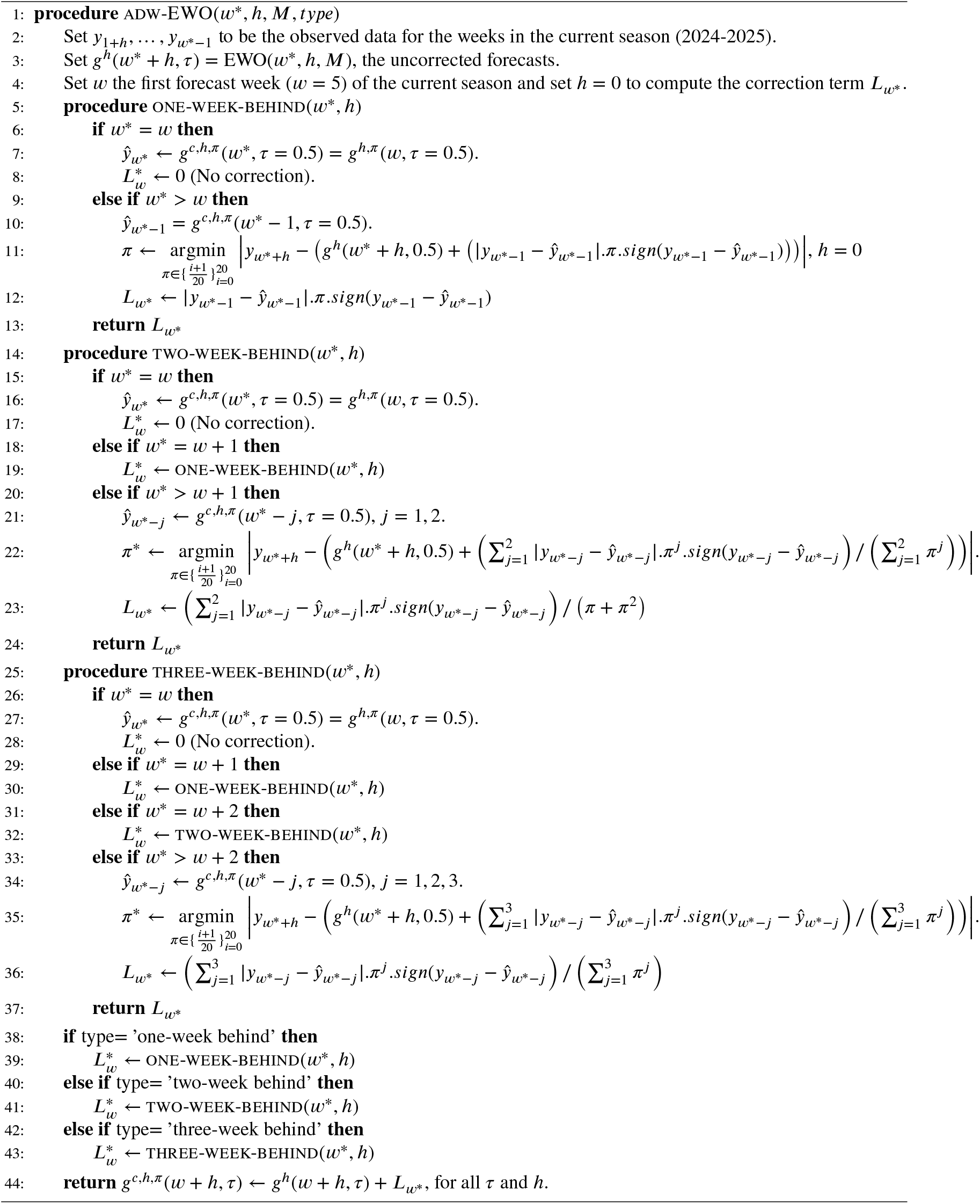

## Behavior of the Discount Factor *π*

**Table 8.**
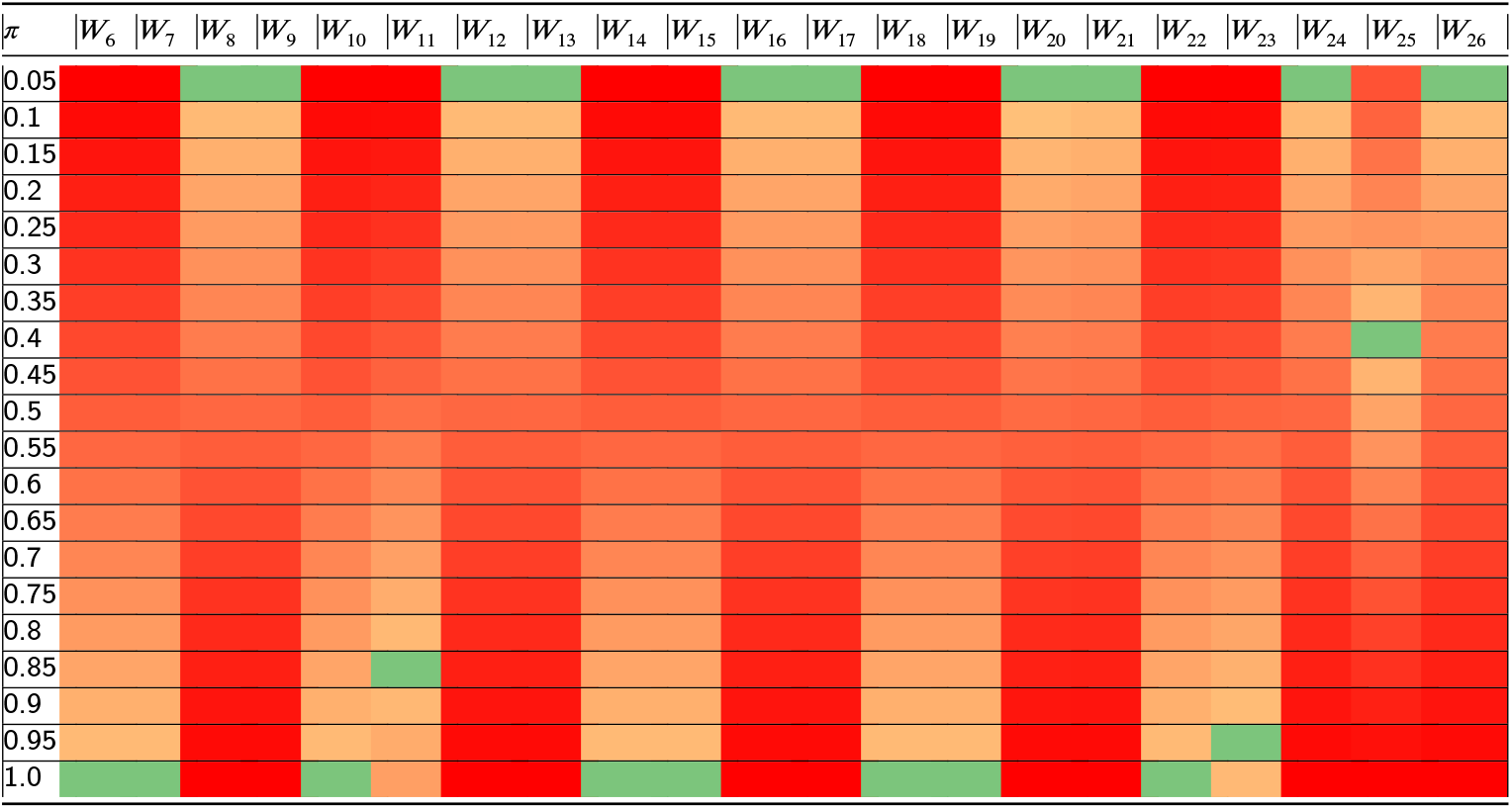
Absolute Errors (AEs) for one-week behind correction with different *π* values. The first column shows *π*, and the remaining columns show the reference weeks *W*_6_ to *W*_26_. Green cells indicate lower AEs, red cells indicate higher AEs.

**Table 9.**
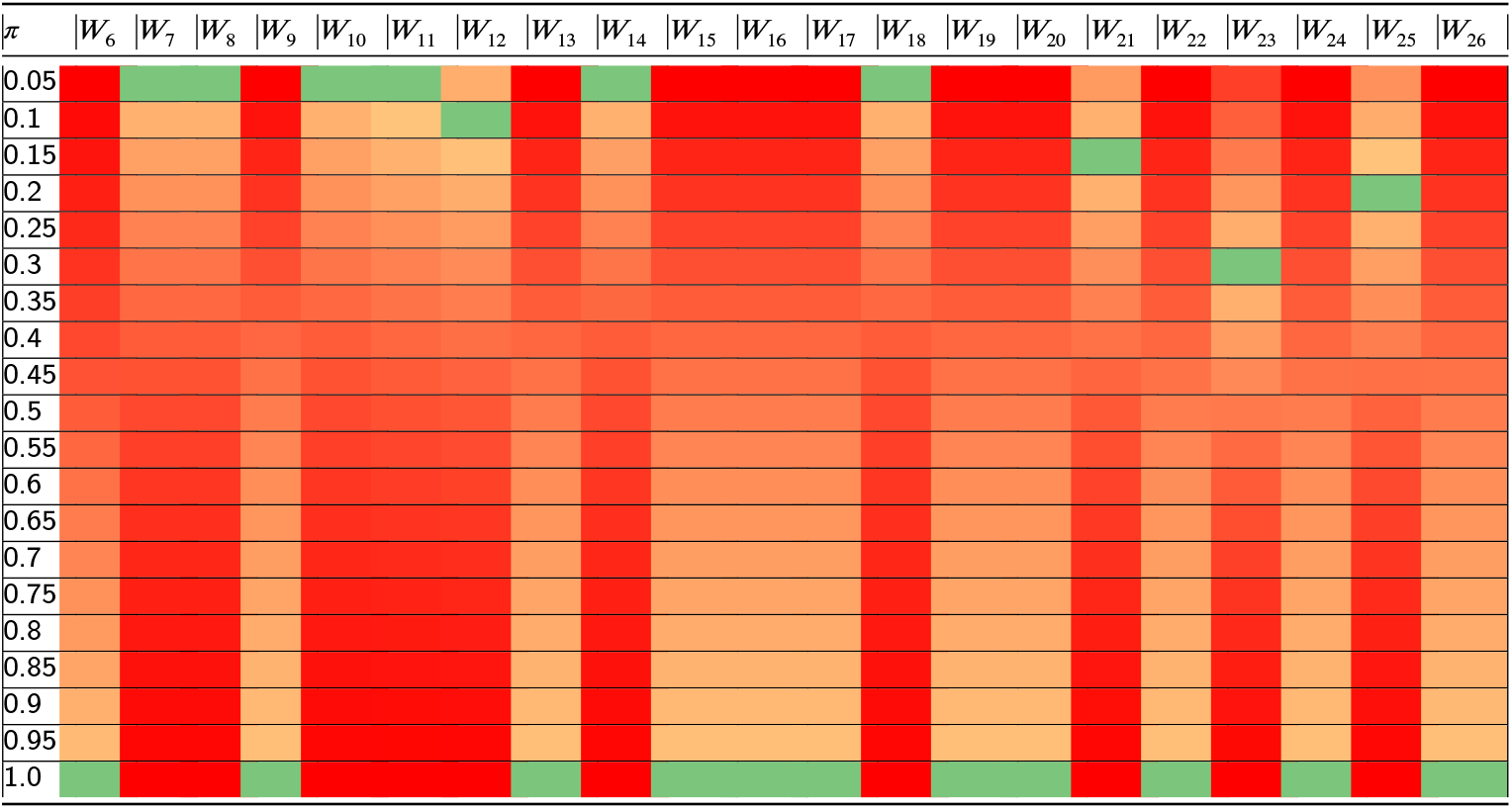
Absolute Errors (AEs) for two-week behind correction with different *π* values. The first column shows *π*, and the remaining columns show the reference weeks *W*_6_ to *W*_26_. Green cells indicate lower AEs, red cells indicate higher AEs..

**Table 10.**
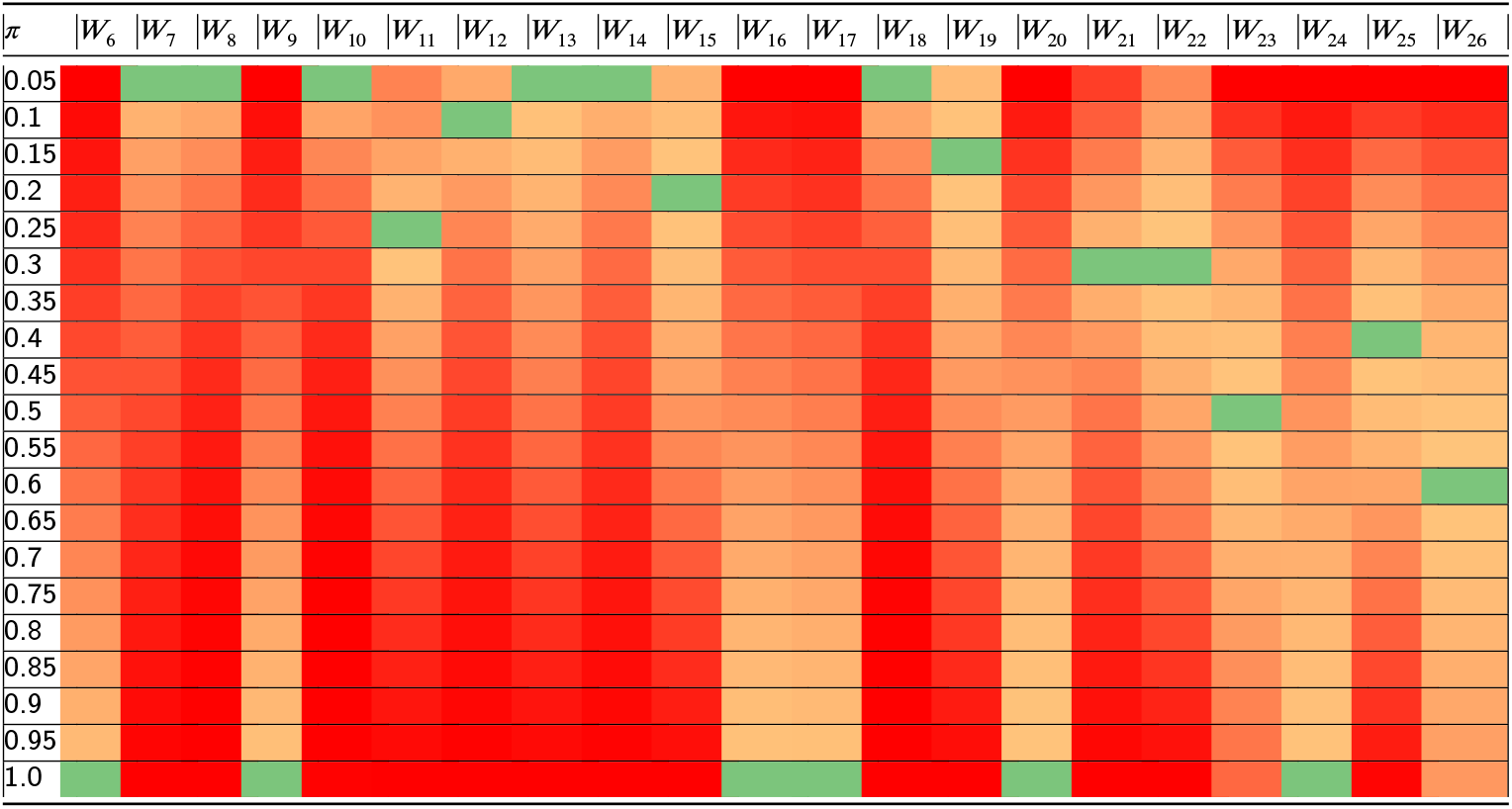
Absolute Errors (AEs) for three-week behind correction with different *π* values. The first column shows *π*, and the remaining columns show the reference weeks *W*_6_ to *W*_26_. Green cells indicate lower AEs, red cells indicate higher AEs.

## Footnotes

1 This represents a series of quantiles starting at 0.025, in increments of 0.05

2 A consistent team is a team that has submitted in all past considered competitions and keeps submitting in the current competition; if a consistent team fails to submit a forecast in a given week *w*, then the baseline model will replace it.

3 For three-week memory, for instance, the formula is 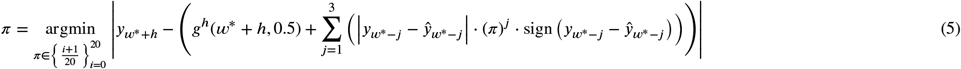

